# Analysis of the number of deaths in Brazil between 2003 and 2020 and possible inferences about the COVID-19 pandemic and history of other diseases

**DOI:** 10.1101/2021.01.23.21250374

**Authors:** Lilian P. Sosman, Andrés R. R. Papa

**Affiliations:** Instituto de Física, UERJ, Rua São Francisco Xavier 524, Maracanã, Rio de Janeiro, 20550-900, RJ, Brazil; Observatório Nacional, Rua General José Cristino 77, São Cristóvão, Rio de Janeiro, 20921-400, RJ, Brazil

**Keywords:** Covid 19, Brazil, National statistics data, epidemics, pandemics

## Abstract

This work explores data on the number of deaths in Brazil since the beginning of the historical series of IBGE, 2003, together with data for the period 2015-2020 of the Transparency Portal. The graphs for total deaths, deaths from violence and deaths in hospitals are discussed. The relationship between them leads to conclusions about the real dimension of the effect of COVID-19 in Brazilian society during the year 2020 and its relative importance to other diseases that had a lesser impact. Hypotheses are also made about the number of deaths in future years.

## INTRODUCTION

Since the first case of COVID-19, officially reported in December 2019 [1], in the city of Wuhan, People’s Republic of China, millions of people have been infected worldwide. Most of these have managed to recover, but unfortunately a significant proportion have died. In February 2020 the Brazilian federal government declared a national emergency [2]. The following month, March 2020, the World Health Organization declared the outbreak a pandemic [3], which was followed by many efforts of all kinds to prevent its spread and minimize its effects.

Brazil registered, as of March 2020, an increasing number of deaths, besides the increase that has always been observed since the beginning of the historical series, registered by IBGE (Brazilian Institute of Geography and Statistics) [4] in 2003, except in 2005.

In this work we study the historical variation of deaths since the beginning of the series, in 2003, to try to evaluate the effect that COVID-19 had on the number of deaths in 2020 and the number that would be expected in the absence of this pandemic. The series, simultaneously, may be useful in assessing epidemics that have affected Brazil over these years. In the work, as auxiliary tools, the numbers of deaths in hospitals and the number of deaths associated to violence throughout Brazil from 2003 to 2019 were also included.

Our study was carried out using public data that are easily accessible to anyone interested and is therefore easy to reproduce. The sources were IBGE [4] and the Civil Registry Transparency Portal [5].

The rest of the work is organized as follows: in the DATA section the databases used for its development are described. In the RESULTS section the graphs on total deaths, violent deaths and hospital deaths from 2003 to 2020 are presented. Data on deaths related to other diseases in the years 2019 and 2020 are also presented. In the DISCUSSION section we analyze the graphs presented in the previous section, as well as the correlations between them, which allow us to arrive at the CONCLUSIONS section where the main findings of our work are summarized.

## DATA

The data used for the number of deaths in Brazil (total, violent and in hospitals) were extracted from IBGE. Data from the Transparency Portal on deaths in Brazil from 2015 to 2020 (the only data available) were also used. Regarding the two databases, it is worth pointing out that in the years that they are included in both databases, the data do not coincide exactly. Data on deaths in Brazil in 2019 and 2020 due to specific diseases were also obtained from the Transparency Portal.

## RESULTS

In figure 1 we can observe the total number of deaths per year in Brazil from 2003 to 2019 (black circles), obtained from IBGE [4] and the total number of deaths per year in Brazil from 2015 to 2019 (black triangles), according to data from the Transparency Portal [5]. Figure 1 also represents the point of intersection of the linear adjustment of these two sets of data for 2020. The black square is the total number of deaths for 2020, obtained from the Transparency Portal [5].

**Figure 1.**
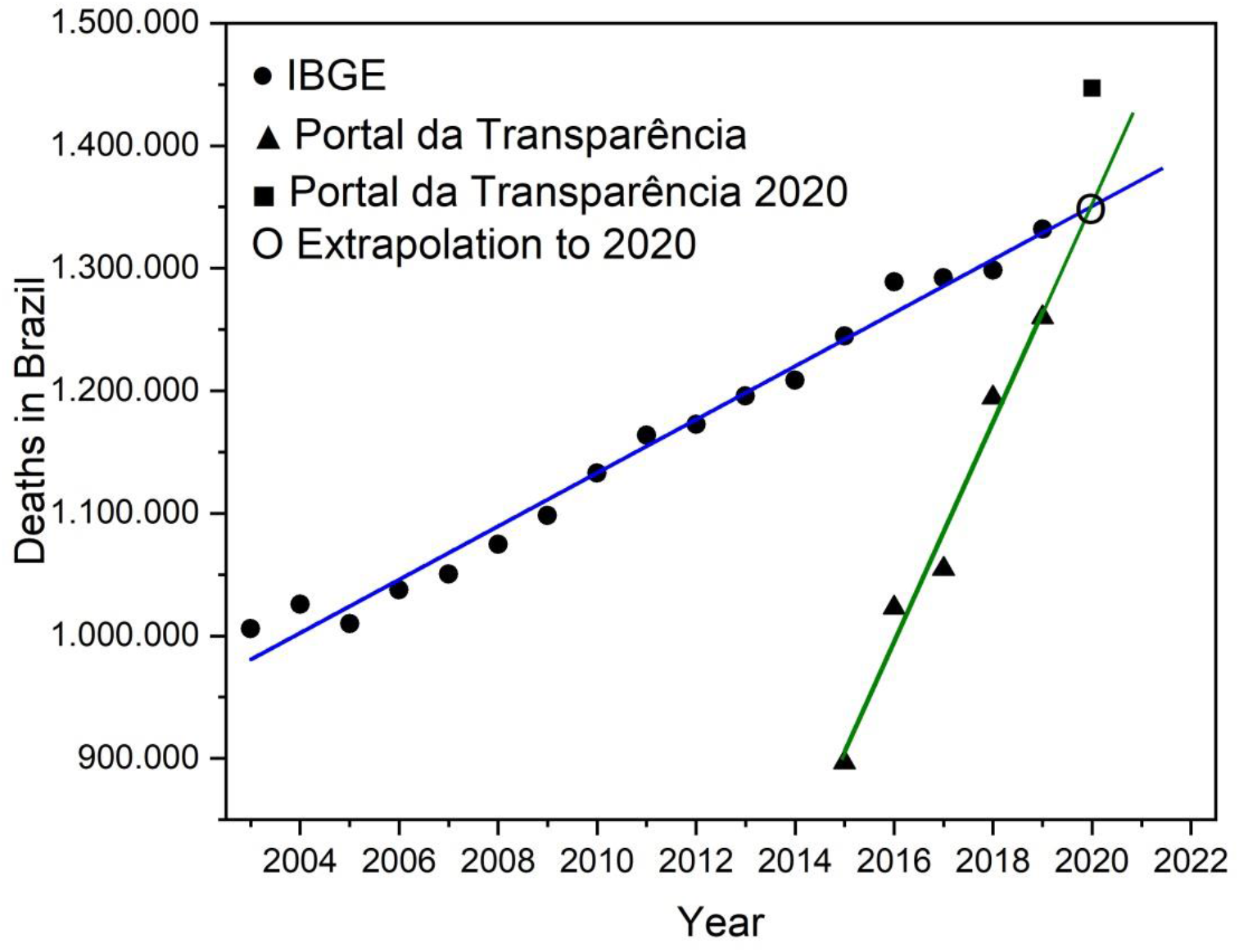
Total number of deaths in Brazil between 2003 and 2019. The black circles correspond to data from IBGE [4] since the beginning of the series in 2003. The blue line is the linear adjustment and these data. The black triangles represent the data of the Transparency Portal available [5]. The green line is the linear adjustment to this data. The point of intersection between the lines corresponds approximately to the extrapolation of both for the year 2020. The black square is the number of deaths in the Transparency Portal for the year 2020. Note that the two sets of data have years in common, but in which the data do not coincide. The Transparency Portal is a more recent base and we believe that it will gradually be completed, so that in the future the slope will be similar to the slope of the linear adjust from IBGE data. Interestingly, in the graph we observe that as of 2020, without a pandemic, the two series would coincide. It is normal that large databases only guarantee their data after a certain time has elapsed. Clear examples of this are the databases of earthquakes and magnetic storms, among others.

The difference between the two points for 2020 (the intersection point of extrapolations and the data from the Transparency Portal for the year 2020) should be observed, which gives the value of 97,287 deaths. If we compare the actual values of deaths for 2019 and 2020, using the data for 2019 from IBGE and the data for 2020 from the Transparency Portal, the difference is 115,042 deaths. If we use the two values from the Transparency Portal, the difference is 187,023 deaths.

It is important to point out at this point that, given that the number of deaths by COVID-19 officially informed on the Transparency Portal [6] until 12/31/2020 was 195,382 [6], the difference between the two values for 2020 (the one extracted from the Transparency Portal and the one obtained by extrapolating the adjustments for the years 2003-2019 and 2015-2019), should be of this order. However, the extrapolation value is approximately half (97,287 deaths) of the registered deaths.

It also draws attention, in IBGE data, to the difference between the years 2015 and 2016 (44,298 deaths), as well as the difference between the years 2009 and 2010 (34,326) and between the years 2010 and 2011 (31,039). Another notable difference occurred between the years 2005 and 2006 (27,452). It is important seen that the only decrease in the number of deaths in the historical series was between two consecutive years, 2004 and 2005 (- 15,839).

Figure 2 shows the number of violent deaths in Brazil between 2003 and 2019. In this graph it is worth noting that between 2009 and 2011 the number remained approximately constant, as well as between 2015 and 2016. Other points that stand out are those between 2017 and 2019 with a systematic drop in the number of violent deaths that led to the lowest number in the historical series in 2019.

**Figure 2.**
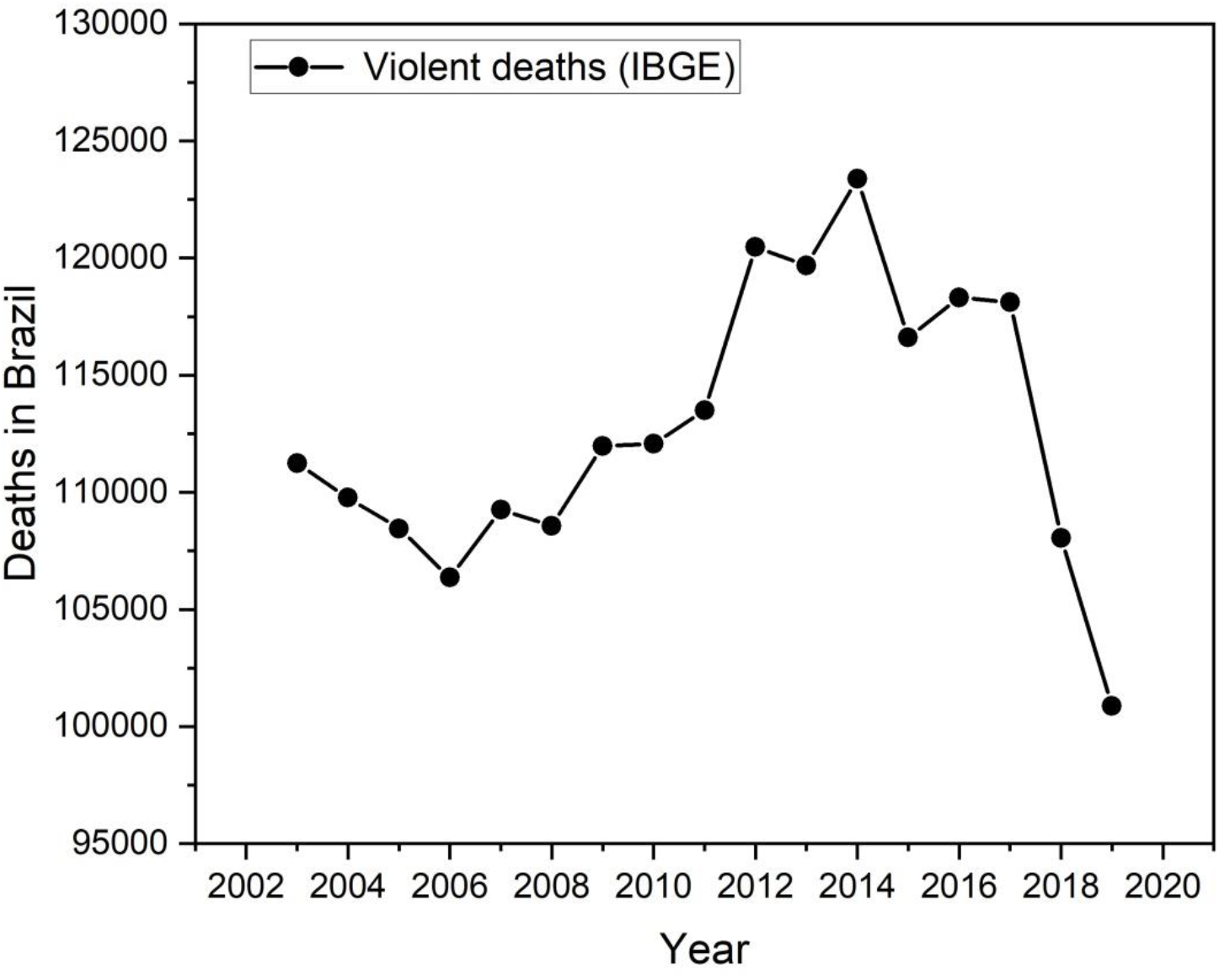
Number of violent deaths in Brazil, between 2003 and 2019. There was a sharp decrease between the years 2017 and 2019, which led to the minimum of the historical series so far.

Figure 3 shows the number of deaths in hospitals from 2003 to 2019. Again, the differences between 2009 and 2010, and between 2015 and 2016, are highlighted. A new jump appeared between the years 2005 and 2006.

**Figure 3.**
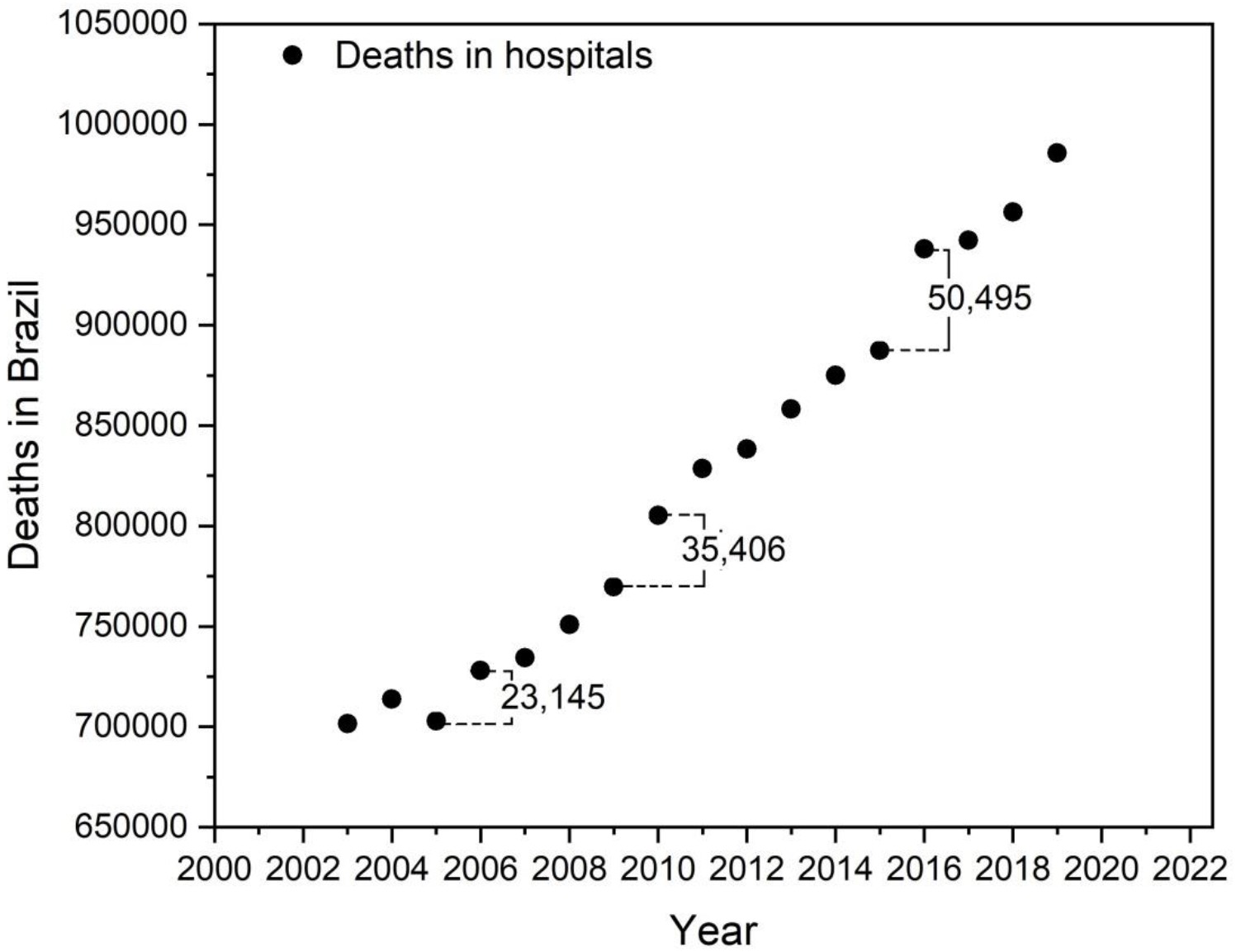
Number of deaths registered in hospitals in Brazil between 2003 and 2019. In this graph, as in figure 1, the differences between the years 2005 and 2006, 2009 and 2010, and 2015 and 2016 are remarkable.

## DISCUSSION

The difference between 2019 and 2020, 77,521 deaths, is the largest ever observed in the historical series (Figure 1). However, observing Figure 2, we can infer that there are two facts that must have impacted this difference. The first is the number of violent deaths in 2019, which is the lowest in the historical series, and which may have led to an increase in the difference in total deaths between the years 2019 and 2020 of the order of 10,000 deaths. The second, which for the time being is partial [7], is the trend towards an increase in the number of violent deaths during the year 2020.

From Figure 1 it can be stated with certainty that COVID-19 brought a significant increase in the number of deaths, far beyond what would be expected by analyzing the historical series [4,5]. However, some characteristics that are highlighted in Figure 1 appear in Figure 3 (deaths in hospitals). From the behavior of the differences between the years 2009 and 2010, and between the years 2010 and 2011, it can be inferred that there was some epidemic that affected the health system in Brazil. We risk that it may have been the first appearance of H1N1 (but this should be the object of more detailed studies). Likewise, the difference between the years 2015 and 2016 may be the signature of another epidemic or epidemics that may have affected Brazil in those years, such as chikungunya, zica and dengue (but as in the case of H1N1, this should be the target of more detailed studies). The difference between 2005 and 2006 could perhaps be related to sporadic appearances of swine fever in that period.

From the comparison of the differences between the years 2010 and 2011, 2015 and 2016 and 2019 and 2020, it can be seen that all of them are of the same order of magnitude. Therefore, it is to be expected that there were a number of false positives, as can be observed by the name adopted in the Transparency Portal - Special Covid-19, in the Registry Panel of summation: “Deaths with suspicion or confirmation of Covid-19”. It is worth mentioning the decrease registered between the years 2019 and 2020 [6] in the number of deaths from other respiratory diseases (not COVID-19) such as pneumonia, which fell by 44 413 deaths [6].

However, another plausible hypothesis is that a large part of the deaths that would occur naturally over 2020 (or even 2021, 2022 and beyond) due to severe comorbidities have been accelerated by the pandemic. If this hypothesis is correct, a number of deaths will be verified in 2021 and 2022, below what is expected from the extrapolation of historical series from 2003 to 2019, shown in Fig. 1. This will obviously occur with the decrease in the number of deaths by COVID-19 by the use of vaccines, treatments and obtaining “herd immunity”. This decrease could be an indication that COVID-19 was definitely removed from our coexistence.

The declaration of COVID-19 as a pandemic brought widespread concern that must have contributed to the increase in the number of deaths by 2020 compared to 2019. One example is the increase in SARS deaths of 15044 [6].

One can mention the abandonment by patients of treatment for pre-existing diseases, suicides and domestic violence, among others. The initial extreme concern must have been caused by the rapid spread of the disease in populations especially vulnerable to it (elderly population in European countries), even in places of high human development. It draws attention to the relatively low morbidity that the virus has caused in populations of places theoretically less prepared to face this type of disorder (Africa). But it is also worth mentioning that the African population is, in general and due to low life expectancy, young. Brazil is not at either extreme.

## CONCLUSIONS

The number of deaths in Brazil in 2020, expected by extrapolation of the historical series 2003-2019 (IBGE) and 2015-2019 (Portal), has increased sharply since the appearance of COVID-19. The total number of deaths, although representing an immeasurable human loss, remained in the same order of magnitude as the number of deaths due, possibly, to other infectious diseases in previous periods. The difference between the total number of deaths in the years 2019 and 2020 may also be associated with an increase in the number of violent deaths in 2020. On the other hand, the number of deaths reported on the Transparency Portal as “Deaths with suspicion or confirmation of COVID-19” far exceeds the statistical projections presented in this work, specifically, the difference between the extrapolation of the historical series of the IBGE and the Portal until 2019, and the data of the Transparency Portal for 2020, as well as seem to be related to the variation in the number of deaths due to other respiratory diseases. It would be very interesting to do similar studies for others countries and eventually to the whole world, to know the actual impact of the COVID-19 worldwide.

## Data Availability

[1] LEI No 13.979, DE 6 DE FEVEREIRO DE 2020, https://www.in.gov.br/en/web/dou/-/lei-n-13.979-de-6-de-fevereiro-de-2020-242078735. Accessed on Jan. 19, 2021.
[2] https://www.gov.br/pt-br/noticias/saude-e-vigilancia-sanitaria/2020/03/oms-classifica-coronavirus-como-pandemia. Accessed on Jan. 19, 2021.
[3] https://sidra.ibge.gov.br/pesquisa/registro-civil/tabelas/brasil/obitos. Tabela 2684. Accessed on Jan. 19, 2021.
[4] https://transparencia.registrocivil.org.br/registros. Accessed on Jan. 19, 2021.
[5] https://transparencia.registrocivil.org.br/especial-covid. Accessed on Jan. 19, 2021.
[6] https://forumseguranca.org.br/anuario-brasileiro-seguranca-publica/. Accessed on Jan. 19, 2021.

## ACKNOWLEDGEMENTS

The authors are grateful to Fundação Carlos Chagas Filho de Amparo à Pesquisa do Estado do Rio de Janeiro (FAPERJ) and Conselho Nacional de Desenvolvimento Científico e Tecnológico (CNPq) for their financial support. L. P. Sosman thanks CNPq for the Research Productivity fellowship.

## BIBLIOGRAPHY

[1] Croda, J. H. R. e Garcia, L. P, Resposta imediata da Vigilância em Saúde à epidemia da COVID-19, Epidemiol. Serv. Saúde 29 (1) 23 Mar 202; https://doi.org/10.5123/S1679-49742020000100021.

[2] LEI Nº 13.979, DE 6 DE FEVEREIRO DE 2020, https://www.in.gov.br/en/web/dou/-/lei-n-13.979-de-6-de-fevereiro-de-2020-242078735. Accessed on Jan. 19, 2021.

[3] https://www.gov.br/pt-br/noticias/saude-e-vigilancia-sanitaria/2020/03/oms-classifica-coronavirus-como-pandemia. Accessed on Jan. 19, 2021.

[4] https://sidra.ibge.gov.br/pesquisa/registro-civil/tabelas/brasil/obitos. Tabela 2684. Accessed on Jan. 19, 2021.

[5] https://transparencia.registrocivil.org.br/registros. Accessed on Jan. 19, 2021.

[6] https://transparencia.registrocivil.org.br/especial-covid. Accessed on Jan. 19, 2021.

[7] https://forumseguranca.org.br/anuario-brasileiro-seguranca-publica/. Accessed on Jan. 19, 2021.

